## Supplementary Information for "Projected health impact of post-discharge malaria chemoprevention among children with severe malarial anaemia in Africa"

Lucy C Okell,[1*] Titus K. Kwambai,[2,3] Aggrey Dhabangi[4], Carole Khairallah[3], Thandile Nkosi-Gondwe[5,6], Robert Opoka[4], Andria Mousa,[1] Melf-Jakob Kühl[7], Tim C. D. Lucas[8], Richard Idro[4], Daniel J. Weiss[9,10], Matthew Cairns[11], Feiko O ter Kuile[3], Kamija Phiri[5,6], Bjarne Robberstad[7], Amani Thomas Mori[*7,12]

1. MRC Centre for Global Infectious Disease Analysis, Department of Infectious Disease Epidemiology, Imperial College, London W2 1PG, UK.
2. Centre for Global Health Research (CGHR), Kenya Medical Research Institute (KEMRI), Kisumu, Kenya
3. Department of Clinical Sciences, Liverpool School of Tropical Medicine (LSTM), Liverpool, United Kingdom
4. College of Health Sciences, Makerere University, Kampala, Uganda.
5. Kamuzu University of Health Sciences, Blantyre, Malawi
6. Training and Research Unit of Excellence, Blantyre, Malawi
7. Section for Ethics and Health Economics, Department of Global Public Health and Primary Care, University of Bergen, P.O. Box 7804, 5020 Bergen, Norway
8. Big Data Institute, University of Oxford, Oxford, UK
9. Malaria Atlas Project, Telethon Kids Institute, Perth Children's Hospital, 15 Hospital Avenue, Nedlands, Australia.
10. Curtin University, Bentley, Australia
11. International Statistics and Epidemiology Group, London School of Hygiene and Tropical Medicine, London, UK.
12. Chr. Michelsen Institute P.O. Box 6033, N-5892 Bergen, Norway.

*Contributed equally, corresponding authors

Lucy Okell, MRC Centre for Global Infectious Disease Analysis, Department of Infectious Disease Epidemiology, Imperial College London, UK.

Amani Thomas Mori, Section for Ethics and Health Economics, Department of Global Public Health and Primary Care, University of Bergen, P.O. Box 7804, 5020 Bergen, Norway.

**Supplementary Methods**

**Post-discharge cohort model**

We developed a compartmental model to describe the incidence of uncomplicated and hospitalised malaria among the trial cohort of children aged 6–59 months during the six months post-discharge follow-up period in PDMC and placebo trial arms. The model has the following states: prophylaxis ($P_{AL}$ and $P_{DP}$), susceptible ($S$), treated uncomplicated malaria ($T_U$), and treated hospitalised malaria ($T_S$) (Figure 4, main text). State transitions are described in the main text and we provide further details here. The equations are below.

Children enter the model into a protected state $P_{AL1}$ on the first day of their treatment with artemether-lumefantrine (AL), which has a gamma-distributed duration with an mean of 13 days.[1] The probability of leaving state $P_{AL1}$ on day $t_d$ after discharge is therefore:

$$r_{pAL}(t_d) = \frac{1}{\Gamma(k_{AL})}\gamma\left(k_{AL}, \frac{t_d}{v_{AL}}\right) - \frac{1}{\Gamma(k_{AL})}\gamma\left(k_{AL}, \frac{t_d - 1}{v_{AL}}\right)$$

where $k_{AL}$ and $v_{AL}$ are the shape and scale parameters of the gamma distribution, respectively. The equation gives the integral of the gamma distribution between the start and end of day $t_d$. The number of children leaving state $P_{AL1}$ each day is therefore: $r_{pAL}(t_d)P_{AL1}(0)$ where all the children are in state $P_{AL1}$ at $t_d = 0$.

Prophylaxis prevents the emergence of blood stage infections from the liver but does not prevent infection with sporozoites, nor liver-stage infection. After the AL prophylaxis wanes, children move to a susceptible state $S$. We assume a constant force of infection, and therefore a constant rate of new blood stage infection, rather than explicitly including a latent period, in line with previous work.[1] That is, children can experience a malaria episode as soon as they transition to the susceptible state.

We model the incidence of both uncomplicated and more severe episodes of malaria requiring hospitalisation in the cohort. The total incidence of these symptomatic episodes occurs at a rate equal to the product of the local entomological inoculation rate (EIR), the probability that an infectious bite leads to infection $b$, the probability of developing symptoms of clinical disease (total uncomplicated and severe) $\phi$ and the relative exposure to mosquito bites among the post-discharge group of children $\xi$ (Table 1, main text).

We further allow that the risk of symptomatic malaria per infectious bite could decline over time since hospital discharge and allow total incidence in the cohort to be scaled by a Weibull survival curve:

$$e^{\left[-\left(\frac{t_d}{\lambda_{risk}}\right)^{\eta_{risk}}\right]}$$

where $t_d$ is the time since hospital discharge and the scale and shape parameters $\lambda_{risk}$ and $\eta_{risk}$, are estimated.

Furthermore, symptomatic malaria incidence is scaled by a similar functional form to that identified in previous analyses[2,3] of the relationship of EIR and the probability of symptomatic malaria:

$$e^{-wE}$$

where the parameter $w$ is estimated. The total incidence of symptomatic malaria in the placebo group $inc_d$ at a given local EIR, and a given time since discharge $t_d$ is therefore:

$$inc_d(t_d) = EIR\ b\phi\xi\, e^{\left[-\left(\frac{t_d}{\lambda_{risk}}\right)^{\eta_{risk}}\right]}\, e^{-wE}$$

Upon developing symptoms, we allowed a two-day delay for treatment seeking ($1/r_{UM}$), after which uncomplicated cases (state $T_U$) receive 13 days of AL post-treatment prophylaxis ($dur_{PAL}$) as described above. This 15-day period was modelled as a fixed duration ($dur_{TU}$), both for computational efficiency and because AL is estimated to have a sharp drop off in protection rather than a gradual decline (Figure S1). We assumed all symptomatic malaria episodes during follow-up were treated with AL in both trial arms, since study clinics were set up and financial reimbursement was given to cover costs of treatment seeking[4] (probability of treatment $f_T = 1$). The drug efficacy, i.e. the probability of successful parasite clearance and protection after AL is denoted $e_{AL}$. Recovered cases return to the *S* state. The proportion of uncomplicated cases who fail treatment ($1 - e_{AL}$) remain in the *S* state and their episode is also counted in the model output. A proportion $\theta$ of the total symptomatic malaria cases require hospitalisation. These enter a treated severe state $T_s$ with a fixed duration of 18 days in total ($dur_{TS}$), which includes 2 days treatment seeking, a mean hospital stay of 3 days ($dur_H$) based on the trial data[4] and 13 days of AL prophylaxis post-discharge.

We track children in the PDMC trial arm separately and denote their states: $S'$, $P'_{AL1}$, $T'_U$, $T'_s$. The model describing these children is the same as the placebo group except for additional PDMC protection against uncomplicated and hospitalised episodes. PDMC is given as three full courses of dihydroartemisinin-piperaquine (DP) starting at the beginning of weeks 2, 6 and 10 post-discharge. Each course contains 3 doses of DP, so that the full PDMC intervention contains 9 doses in total. The efficacy of DP is denoted $e_{DP}$. We modelled DP prophylaxis as a probability of prevention of reinfection, $p_{pDP}$ that declines over time since treatment, using a Weibull-survival function with a mean of 28 days based on previous fitting to clinical trial data:[5]

$$p_{pDP}(t_{DP}) = p_{ad} e_{DP} e^{\left[-\left(\frac{t_{PMC}}{\lambda_{DP}}\right)^{\eta_{DP}}\right]}$$

where $t_{DP}$ is the time since the last PDMC treatment, $p_{ad}$ is the probability of adhering to the current course of DP while $\lambda_{DP}$ and $\eta_{DP}$ represent the scale and shape parameters of the Weibull distribution. The efficacy of DP is high (99%) and adherence during the trial is also high (97.3-99.0%). During model fitting we therefore chose to simply reduce the probability of protection proportionately to the efficacy and adherence (as shown in the equation above) for computational efficiency, rather than separately track children who do not receive protection from one of the DP courses. We allowed for different adherence to each of the 3 courses of PDMC $p_{ad1}$, $p_{ad2}$, $p_{ad3}$. We incorporated the measured percentage of children adhering to treatment in the trial setting[4] when fitting to the trial data (Table 1, main text). For simplicity, we assumed that for each course of PDMC, caregivers either gave all 3 doses or none, which was relatively consistent with observations during implementation (only 1.5% of children received 1-2 doses of DP per treatment course, with the remainder taking all or none).

We further explored whether the probability of a symptomatic malaria episode being severe enough to require hospitalisation, $\theta$, could be different in children with active PDMC drug protection ($\theta_2$) compared to children without ($\theta_1$). We defined active drug protection as inhibitory drug levels providing >1% probability of protection.

In the model, we incorporated the number of children recruited in each trial site to each trial arm, and the loss to follow up in each group. At the time a child was lost to follow up (due to leaving the study, or dying) we reduced the total population accordingly in all model states in that trial site and arm. E.g. if there were 100 children on day $t_d - 1$, and 99 on day $t_d$, we multiply the numbers in all model states on day $t_d - 1$ by 0.99 to obtain the numbers still present on day $t_d$.

The model was written in discrete time using daily time steps, with $t_d$ being the time since hospital discharge. The cohort model is run for 25 weeks to match the follow up duration in the data. All rates were converted to daily transition probabilities using the equation:

$$y = 1 - e^{-r}$$

where $y$ is the transition probability and $r$ is the daily rate.[6] The incidence of all symptomatic episodes per day, $inc_d$, was converted to the probability of an episode by the end of the day $y_d$.

The full system of equations for the placebo group is as follows:

$$P_{AL1}(t_d) = P_{AL1}(t_d - 1) - r_{pAL}(t_d - 1)P_{AL1}(0)$$

$$\begin{aligned} S(t_d) = S(t_d - 1) + r_{pAL}(t_d - 1)P_{AL1}(0) - y_d(t_d - 1)e_{AL}S(t_d - 1) \\ + y_d(t_d - dur_{TU})e_{AL}S(t_d - dur_{TU})(1 - \theta_1) \\ + y_d(t_d - dur_{TS})e_{AL}S(t_d - dur_{TS})\theta_1 \end{aligned}$$

$$\begin{aligned} T_U(t_d) = T_U(t_d - 1) + y_d(t_d - 1)e_{AL}S(t_d - 1)(1 - \theta_1) \\ - y_d(t_d - dur_{TU})e_{AL}S(t_d - dur_{TU})(1 - \theta_1) \end{aligned}$$

$$T_S(t_d) = T_S(t_d - 1) + y_d(t_d - 1)e_{AL}S(t_d - 1)\theta_1 - y_d(t_d - dur_{TS})e_{AL}S(t_d - dur_{TS})\theta_1$$

The system of equations for the PDMC intervention group is:

$$P'_{AL}\ (t_d) = P'_{AL1}(t_d - 1) - r_{pAL}(t_d - 1)P'_{AL}\ (0)$$

$$\begin{aligned} S'(t_d) = S'(t_d - 1) + r_{pAL}(t_d - 1)P'_{AL1}(0) - y_d(t_d - 1)e_{AL}S'(t_d - 1)(1 - p_{DP}(t_{DP})p_{ad}) \\ + y_d(t_d - dur_{TU})e_{AL}S'(t_d - dur_{TU})(1 - \theta)\left(1 - p_{pDP}(t_{DP})p_{ad}\right) \\ + y_d(t_d - dur_{TS})e_{AL}S'(t_d - dur_{TS})\theta\left(1 - p_{pDP}(t_{DP})p_{ad}\right) \end{aligned}$$

$$\begin{aligned} T'_U(t_d) = T'_U(t_d - 1) + y_d(t_d - 1)e_{AL}S'(t_d - 1)(1 - \theta)\left(1 - p_{pDP}(t_{DP})p_{ad}\right) \\ - y_d(t_d - dur_{TU})e_{AL}S'(t_d - dur_{TU})(1 - \theta)\left(1 - p_{pDP}(t_{DP})p_{ad}\right) \end{aligned}$$

$$\begin{aligned} T'_S(t_d) = T'_S(t_d - 1) + y_d(t_d - 1)e_{AL}S'(t_d - dur_{TS})\theta\left(1 - p_{pDP}(t_{DP})p_{ad}\right) \\ - y_d(t_d - 18)e_{AL}S'(t_d - dur_{TS})\theta\left(1 - p_{pDP}(t_{DP})p_{ad}\right) \end{aligned}$$

At $t_d = 0$, all individuals in the placebo arm start in state $P_{AL}$ and all individuals in the PDMC arm start in state $P'_{AL1}$. The protection from PDMC, $p_{pDP}$, is 0 before day 14. The time since PDMC, $t_{DP}$, is $t_d - 14$ between follow up days 14-42, $t_d - 42$ between day 42-70, and $t_d - 70$ thereafter. The proportions of children adhering to PDMC $p_{ad}$ during these 3 time periods are $p_{ad}$ , $p_{ad}$ and $p_{ad}$ , respectively (Table S1). The probability of a symptomatic episode being more severe and requiring hospitalisation, $\theta$, is $\theta_2$ when the child is protected by DP with probability of protection $p_{pDP} > 0.01$ and $\theta_1$ when $p_{pDP} \leq 0.01$.

### Model fitting and validation

We obtained prior estimates for the EIR in the areas surrounding each hospital in the post-discharge studies. Since EIR is not a commonly measured metric, we generated modelled EIR from the average of parasite prevalence estimates in 2-10 year olds within a 20 km radius of each hospital using our pre-existing transmission model[23,25]. These parasite prevalence estimates are from the Malaria Atlas Project (MAP) global map averaged over the years 2016-2018, when the trial was conducted. We used these EIR estimates as semi-informative prior medians, assuming a gamma distribution with 10% coefficient of variation (Table S1).

As described above, we estimated the total incidence of uncomplicated plus hospitalised malaria episodes as the product of the local EIR, the probability that an infectious bite leads to infection $b$, the probability of symptoms (total uncomplicated and severe) $\phi$ and the relative exposure to mosquito bites among the post-discharge group of children $\xi$ (Table 1). Children usually experience lower exposure to bites than adults simply due to having lower body surface area. [23,24] However, it is unknown whether exposure among the post-discharge group is higher or lower than among other children of the same age in the general population. During the initial analysis we noticed that the maximum incidence of symptomatic malaria in the post-discharge children could be slightly higher than the expected average EIR (see also Figure S6), and hence we introduced the term $\xi$ allowing a different exposure in this group of children. We set the allowed range for $b\phi\xi$ between 0 and 2, so that the maximum incidence could not be more than 2-fold higher than the average EIR.

We also estimated the unknown parameters describing the relationship between the risk of an episode per infectious bite, EIR and time post-discharge described above ($\lambda_{risk}$, $\eta_{risk}$, $w$) using uninformative priors (Table S1).[7] The parameters describing the proportion of episodes requiring hospitalisation in children without PDMC and with active PDMC protection (see above), $\theta$, were given a uniform prior between 0-1.

We fit to incidence data from weeks 1-25 of follow up and excluded the final week of follow up (week 26) due to anomalous results. The recorded incidence of uncomplicated malaria was much higher in the last week of follow up than in any other week and was a clear outlier (Figure S7). This was driven by cases from the trial site in Jinja only. Model fitting was undertaken using MCMC in the RStan software.[8]

**Population modelling of PDMC demand and impact in different epidemiological settings**

To estimate PDMC demand and impact in different geographic areas, we combined information from the following databases and models. Modelled estimates were used due to the lack of complete primary country-level data.

1) Incidence of hospitalised SMA in the general population of 3 month -9 year olds in relation to parasite prevalence, as modelled by Paton *et al*[9] based on data from Kenya, Tanzania and Uganda.
2) Parasite prevalence in 2-10 year olds in country subnational regions from the Malaria Atlas Project.[10,11]
3) The incidence of uncomplicated and hospitalised malaria in 0-5 year olds in the 6 months after discharge from hospital following a severe anaemia episode, in relation to EIR, from our current analysis of the Kwambai *et al*[4] study (Figures 2 & 3, main text).
4) A well-established transmission model[2,12,13] (Imperial College London; 'IC model'). This was used to translate between key metrics in the above model inputs: (a) EIR and prevalence in 2-10 year olds, (b) the incidence of SMA in 0-5 year olds relative to 0-10 year olds. This model has been calibrated against a wide range of epidemiological data,[3] including hospitalised malaria incidence in different transmission settings and age groups.[12] The hospitalised malaria component was previously fitted to incidence data from nine sites in different African

countries where populations were considered to have good transport networks to hospital and lived a maximum of 19 km away.[12] The model was also calibrated to hospitalised SMA data.

We constructed a population model tracking SMA in all under five year olds which incorporated the different sources of information 1-4 (Figure S2). The post-discharge cohort as described above was embedded within this model, and we also allowed for additional detail and factors that are different outside the trial setting. These include the possibility of not going to hospital and not receiving effective treatment for uncomplicated malaria. The model also allows that SMA increases risk of future SMA in an iterative process, which results in episodes being more concentrated in particular individuals. We stratify the population of under five year olds into 'low-risk': those who have not experienced an SMA episode in the last 6 months, and 'high-risk': those who have had SMA and are at higher risk of subsequent malaria episodes. We now refer to the latter group as 'post-SMA' rather than 'post-discharge', since they may not all have accessed hospital care for the original episode. We further stratify the model into groups depending on the different number of PDMC doses received. We assume that the high-risk post SMA group is described well by the post-discharge cohort model above. That model was calibrated to data on severe anaemia patients from the trial[4] who did not all have malaria, however this group without malaria was a minority (15% of the cohort).

Children transition through the model as follows. Those in the low-risk group, i.e. who have not experienced SMA in the past 6 months, are in state $G$ and new SMA episodes occur in this group at rate $inc_{gSMA}$. SMA cases have a probability of accessing hospital care, $p_H$. Those who are hospitalised enter state $H$ with a fixed 3-day stay in hospital and have a probability of dying, $q_{SMAH}$. Those who survive then follow a near-identical process to those in the post-discharge cohort model described earlier. We assume that all hospitalised individuals receive AL at discharge as per standard severe malaria guidelines and we model this as a fixed 13-day period of protection in state $P_{AL}$ . Children who do not receive PDMC follow the same path as those in the post-discharge cohort model placebo group described previously. Children who receive PDMC but do not then take the first dose enter state $S_H$ (susceptible, previously received PDMC). That is, they experience new symptomatic malaria episodes at rate $inc_d$, dependent on EIR and time since discharge. The trial settings only included areas with transmission intensity up to EIR of around 30 per year (Figure 3, main text). Since the relationship between EIR and post-discharge incidence of malaria is unknown for higher EIRs, we assumed that post-discharge incidence would plateau at EIR=30 and not increase further nor decrease in higher transmission settings (only 3.8% of regions modelled have an estimated EIR>30). During these episodes children have a probability of requiring hospitalisation $\theta$ and a probability $1-\theta$ of having uncomplicated malaria that only requires an outpatient visit. In contrast to the trial setting, we assume the proportion of children receiving treatment for uncomplicated episodes $f_T$, is <100%. Likewise, of those who would have been hospitalised in the trial, the proportion $p_H$ who access hospital care now is <100%. Given lack of data on the probability of accessing hospital care, we assume $p_H$ is the same for lower risk and higher risk children, and for different types of severe malaria. We vary the assumed value of $p_H$ from 30-70% based on the recent CARAMAL study tracking children with suspected severe malaria in community settings.[14] We also assume that the probability of hospitalisation is not related to previous hospitalisation status. If children access hospital care they enter state $T_S$ as above, or if they receive AL for uncomplicated malaria they enter state $T_U$. After fixed durations they return to the susceptible $S_H$ state.

In this population model, episodes of SMA occur at many time points and children enter the post-SMA state at different times, unlike in the cohort model described above. In order to track the time since SMA for the purpose of modelling changing risk, we separated the children in the high-risk groups into daily compartments from week 3 (when they enter the susceptible state after AL protection ends) up to 25 weeks post-SMA, resulting in 161 high risk days ($n_{HR}$). We assume the percentage of episodes requiring hospitalisation which have SMA during this time ($p_{SMA}$) is the same as during the trial[4] (SMA episodes / total hospitalised malaria episodes = 44%). Those who experience recurrent SMA during the high-risk period once again have probability of hospitalisation $p_H$ and subsequently return to the beginning of the high-risk period. After 25 weeks, children return to the low-risk *G* state if they have not experienced any more SMA episodes.

PDMC is given to hospitalised children at discharge with probability $z_{PDMC}$ (i.e. PDMC coverage). The probabilities of adherence to each of the 3 PDMC courses on days 14, 42 and 70 post-discharge are $p_{ad1}$, $p_{ad2}$ and $p_{ad3}$. We assumed the same adherence as observed during a recent PDMC implementation trial in Malawi,[15] where all 3 PDMC courses were given to caregivers at the time of discharge from hospital without further adherence reminders (Table 1, main text). Children could therefore miss a PDMC course, for example, taking PDMC course 1 at day 14 and course 3 at day 70, but missing the day 42 course. However as above, we assume they do not take partial PDMC courses (i.e. they always either take the full course of 3 doses, or none). Children receiving PDMC have a probability of protection against reinfection provided by DP which changes by day since treatment according to the Weibull survival curve described earlier. We stratify the model according to the different PDMC courses that children may receive. For example, children enter state $PDMC_1$ if they get the first course or remain in the susceptible high-risk state $S_H$ if not. From these states, they then transition to state $PDMC_2$ only if they receive the second PDMC course, and state $PDMC_3$ only if they receive the third course. For example, children who only received the first course would remain in state $PDMC_1$, where the probability of protection declines to zero over time, so that children in this state eventually acquire the same risk as those with no PDMC in state $S_H$ (at the equivalent time after the original SMA episode). The probability of protection at time *j* since discharge in states $PDMC_1$, $PDMC_2$, and $PDMC_3$ is denoted $p_{pDP}$ , $p_{pDP2}$ and $p_{pDP3}$.

A proportion of SMA cases $(1 - p_H)$ do not access hospital care and they enter state $D_C$ (disease in the community). Those who survive transition from $D_C$ to the susceptible high-risk post-SMA state $S_C$ (susceptible, community, did not receive PDMC) at the same time that those who were hospitalised enter the susceptible state $S_H$ post-discharge. Little is known about disease progression outside hospital. Due to lack of data, we do not model any recurrent malaria episodes in state $D_C$. This assumption has no effect on the comparison of the modelled scenarios with and without PDMC intervention since SMA cases outside hospital would be unaffected by PDMC. Children who are hospitalised with SMA and do not receive PDMC also enter state $S_C$ after their period of AL protection is ended.

The full system of equations for the population model at time *t* is below. It was coded using discrete daily time steps to be consistent with the post discharge cohort model. All children begin in state *G* at time 0 and then the model is run to equilibrium. The high risk, post-SMA states $S_H$, $S_C$, $PDMC_1$, $PDMC_2$, $PDMC_3$, $T_S$, $T_U$ are 2D arrays containing the number of individuals in the state at time *t* and day $j$.

The total SMA cases at time *t* is:

$$C_{SMA}(t) = y_{gSMA}G(t-1) + \sum_{j=1}^{n_{HR}} y_d(j)p_{SMA}\left[\theta_1 S_H(t-1,j) + \theta_1 S_C(t-1,j) + \sum_{k=1}^{3}\left(1 - p_{pDPk}(j)\right)\theta PDMC_k(t-1,j)\right]$$

where $\theta$ is the risk of developing disease requiring hospitalisation among symptomatic cases and is equal to $\theta_1$ or $\theta_2$ depending whether the probability of protection from DP is currently less than or more than 0.01 in each PDMC state, respectively. $y_{gSMA}$ is the risk of having a new SMA episode per day in the G state, calculated from $inc_{gSMA}$ in the same way that $y_d$ was calculated from $inc_d$ above.

The total number of other malaria cases without severe anaemia who require hospitalisation in the high-risk group at time *t* and on day *j* is then similarly:

$$C_S(t,j) = y_d(j)(1-p_{SMA})\left[\theta_1 S_H(t,j) + \theta_1 S_C(t,j) + \sum_{k=1}^{3}\left(1-p_{pDPk}(j)\right)\theta PDMC_k(t,j)\right]$$

We do not track non-SMA severe cases in the general population, since we are specifically interested in the effect of SMA in increasing future risk, and there is no specific evidence that other types of severe malaria do so. The equations for the state variables are:

$$G(t) = G(t-1) - y_{gSMA}G(t-1) + \left(1-y_d(n_{HR})\right)\left(S_H(t-1,n_{HR}) + S_C(t-1,n_{HR}) + PDMC_1(t-1,n_{HR}) + PDMC_2(t-1,n_{HR}) + PDMC_3(t-1,n_{HR})\right) + T_U(t-1,n_{HR}) + T_S(t-,n_{HR}) + +p_H\left(q_{SMAH}C_{SMA}(t-1) + q_H C_S(t-1)\right) + (1-p_H)\left(q_{SMAC}C_{SMA}(t-1) + q_C C_S(t-1)\right)$$

$$H(t) = H(t-1) + p_H(1-q_{SMAH})C_{SMA}(t-1) - p_H(1-q_{SMAH})C_{SMA}(t-dur_H)$$

$$D_C = D_C(t-1) + (1-p_H)(1-q_{SMAC})C_{SMA}(t-1) - (1-p_H)(1-q_{SMAC})C_{SMA}(t-dur_{TS})$$

$$P_{AL1}(t) = P_{AL1}(t-1) + p_H(1-q_{SMAH})C_{SMA}(t-dur_H) - p_H(1-q_{SMAH})C_{SMA}(t-dur_{TS})$$

For *j=0:*

$$S_H(t,0) = p_H(1-q_{SMAH})z_{PDMC}(1-p_{ad1})C_{SMA}(t-dur_{TS})$$

$$S_C(t,0) = [p_H(1-q_{SMAH})(1-z_{PDMC}) + (1-p_H)(1-q_{SMAC})]C_{SMA}(t-dur_{TS})$$

$$PDMC_1(t,0) = p_H(1-q_{SMAH})z_{PDMC}p_{ad1}C_{SMA}(t-dur_{TS})$$

$$PDMC_2(t,0) = PDMC_3(t,0) = T_U(t,0) = T_S(t,0) = 0$$

The state variables at time t and on day j since entering the high-risk state are as follows. We use indicator variables $i_j$ which are 1 on day j and 0 otherwise. $i_{PM}$ and $i_{PMC3}$ are indicator variables that are 1 on the days that the 2nd or 3rd course of PDMC begins, and 0 otherwise, and $p_{ad}$ is $p_{ad1}$, $p_{ad2}$and $p_{ad}$ for the 1st, 2nd and 3rd courses of PDMC.

For *j>0:*

$$S_C(t,j) = S_C(t-1,j-1) - y_d(j-1)\theta_1 p_{SMA}(t-1)S_C(t-1,j-1)$$
$$-y_d(j-1)(1-\theta_1)f_T e_{AL} S_C(t-1,j-1)$$
$$-y_d(j-1)\theta_1(1-p_{SMA})[p_H + (1-p_H)(1-q_C)f_T e_{AL} + (1-p_H)q_C]S_C(t-1,j-1)$$
$$+y_d(j-dur_{TU})(1-\theta_1)f_T e_{AL} S_C(t-dur_{TU}, j-dur_{TU})$$
$$+y_d(j-dur_{TS})\theta_1(1-p_{SMA})p_H(1-q_H)S_C(t-dur_{TS}, j-dur_{TS})$$
$$+y_d(j-dur_{TU})\theta_1(1-p_{SMA})(1-p_H)(1-q_C)f_T e_{AL} S_C(t-dur_{TU}, j-dur_{TU})$$

$$S_H(t,j) = S_H(t-1,j-1) - y_d(j-1)\theta_1 p_{SMA}(t-1)S_H(t-1,j-1)$$

$$-y_d(j-1)(1-\theta_1)f_T e_{AL} S_H(t-1,j-1)$$

$$-y_d(j-1)\theta_1(1-p_{SMA})S_H(t-1,j-1)[p_H + (1-p_H)(1-q_C)f_T e_{AL} + (1-p_H)q_C]$$

$$+y_d(j-dur_{TU})(1-\theta_1)f_T e_{AL} S_H(t-dur_{TU}, j-dur_{TU})$$

$$+y_d(j-dur_{TS})\theta_1(1-p_{SMA})p_H(1-q_H)S_H(t-dur_{TS}, j-dur_{TS})$$

$$+y_d(j-dur_{TU})\theta_1(1-p_{SMA})(1-p_H)(1-q_C)f_T e_{AL} S_H(t-dur_{TU}, j-dur_{TU})$$

$$-i_{PMC2}p_{ad}\ S_H(t-1,j-1)$$

$$-i_{PM}\quad p_{ad}\ S_H(t-1,j-1)$$

$$PDMC_1(t,j) = PDMC_1(t-1,j-1)$$

$$- y_d(j-1)\theta p_{SMA}(t-1)PDMC_1(t-1,j-1)\left(1-p_{pDP1}(j-1)\right)$$

$$-y_d(j-1)(1-\theta)f_T e_{AL} PDMC_1(t-1,j-1)\left(1-p_{pDP1}(j-1)\right)$$

$$-y_d(j-1)\theta(1-p_{SMA})PDMC_1(t-1,j-1)\left(1-p_{pDP1}(j-1)\right)[p_H + (1-p_H)(1-q_C)f_T e_{AL}$$

$$+ (1-p_H)q_C]$$

$$+y_d(j-dur_{TU})(1-\theta)f_T e_{AL} PDMC_1(t-dur_{TU}, j-dur_{TU})\left(1-p_{pDP1}(j-dur_{TU})\right)$$

$$+y_d(j-dur_{TS})\theta(1-p_{SMA})p_H(1-q_H)PDMC_1(t-dur_{TS}, j-dur_{TS})\left(1-p_{pDP1}(j-dur_{TS})\right)$$

$$+y_d(j-dur_{TU})\theta(1-p_{SMA})(1-p_H)(1-q_C)f_T e_{AL} PDMC_1(t-dur_{TU}, j-dur_{TU})\Big(1$$

$$- p_{pDP1}(j-dur_{TU})\Big)$$

$$-i_{PMC2}p_{ad2}PDMC_1(t-1,j-1)$$

$$-i_{PMC3}p_{ad3}PDMC_1(t-1,j-1)$$

$$PDMC_2(t,j) = PDMC_2(t-1,j-1) + i_{PM}\quad p_{ad2}\left(PDMC_1(t-1,j-1) + S_H(t-1,j-1)\right)$$

$$- y_d(j-1)\theta p_{SMA}(t-1)PDMC_2(t-1,j-1)\left(1-p_{pD}\quad(j-1)\right)$$

$$-y_d(j-1)(1-\theta)f_T e_{AL} PDMC_2(t-1,j-1)\left(1-p_{pDP2}(j-1)\right)$$

$$-y_d(j-1)\theta(1-p_{SMA})PDMC_2(t-1,j-1)\left(1-p_{pDP2}(j-1)\right)[p_H + (1-p_H)(1-q_C)f_T e_{AL}$$

$$+ (1-p_H)q_C]$$

$$+y_d(j-dur_{TU})(1-\theta)f_T e_{AL} PDMC_2(t-dur_{TU}, j-dur_{TU})\left(1-p_{pDP2}(j-dur_{TU})\right)$$

$$+y_d(j-dur_{TS})\theta(1-p_{SMA})p_H(1-q_H)PDMC_2(t-dur_{TS}, j-dur_{TS})\left(1-p_{pDP2}(j-dur_{TS})\right)$$

$$+y_d(j-dur_{TU})\theta(1-p_{SMA})(1-p_H)(1-q_C)f_T e_{AL} PDMC_2(t-dur_{TU}, j-dur_{TU})\Big(1$$

$$- p_{pDP2}(j-dur_{TU})\Big)$$

$$-i_{PMC3}p_{ad3}PDMC_2(t-1,j-1)$$

$$PDMC_3(t,j) = PDMC_3(t-1,j-1) + i_{PMC3}p_{ad3}\big(S_H(t-1,j-1) + PDMC_1(t-1,j-1) + PDMC_2(t-1,j-1)\big) - y_d(j-1)\theta p_{SMA}(t-1)PDMC_3(t-1,j-1)\left(1-p_{pDP3}(j-1)\right)$$

$$-y_d(j-1)(1-\theta)f_T e_{AL}PDMC_3(t-1,j-1)\left(1-p_{pDP3}(j-1)\right)$$

$$-y_d(j-1)\theta(1-p_{SMA})PDMC_3(t-1,j-1)\left(1-p_{pDP3}(j-1)\right)[p_H + (1-p_H)(1-q_C)f_T e_{AL} + (1-p_H)q_C]$$

$$+y_d(j-dur_{TU})(1-\theta)f_T e_{AL}PDMC_3(t-dur_{TU},j-dur_{TU})\left(1-p_{pDP3}(j-dur_{TU})\right)$$

$$+y_d(j-dur_{TS})\theta(1-p_{SMA})p_H(1-q_H)PDMC_3(t-dur_{TS},j-dur_{TS})\left(1-p_{pDP3}(j-dur_{TS})\right)$$

$$+y_d(j-dur_{TU})\theta(1-p_{SMA})(1-p_H)(1-q_C)f_T e_{AL}PDMC_3(t-dur_{TU},j-dur_{TU})\left(1 - p_{pDP3}(j-dur_{TU})\right)$$

$$T_U(t,j) = T_U(t-1,j-1)$$

$$+y_d(j-1)(1-\theta)f_T e_{AL}\left[S_H(t-1,j-1) + S_C(t-1,j-1) + \sum_{k=1}^{3}\left(1-p_{pDPk}(j-1)\right)PDMC_k(t-1,j-1)\right]$$

$$-y_d(j-dur_{TU})(1-\theta)f_T e_{AL}\left[S_H(t-dur_{TU},j-dur_{TU}) + S_C(t-dur_{TU},j-dur_{TU}) + \sum_{k=1}^{3}\left(1-p_{pDPk}(j-dur_{TU})\right)PDMC_k(t-dur_{TU},j-dur_{TU})\right]$$

$$T_S(t,j) = T_S(t-1,j-1)$$

$$+(p_H(1-q_H) + (1-p_H)(1-q_c)f_T e_{AL})C_S(t-1,j-1)$$

$$-p_H(1-q_H)C_S(t-dur_{TS},j-dur_{TS})$$

$$-(1-p_H)(1-q_c)f_T e_{AL}C_S(t-dur_{TU},j-dur_{TU})$$

**Population model calibration and simulation**

We calibrated the model of SMA in the whole population to different epidemiological settings as follows. For each subnational region of Africa, we obtained the parasite prevalence in 2-10 year olds from the Malaria Atlas project 2019 map,[10,11] and from this, estimated EIR using the IC transmission model.[2] We estimated the total incidence of hospitalised SMA for each area in children aged 3 months-9 years using the recent relationship with parasite prevalence published by Paton *et al.*[9] To convert this to incidence of hospitalised SMA in 0-5 year olds, we generated the ratio of SMA incidence in 3 month-9 year olds to 0-5 year olds in each parasite prevalence category (binned to the nearest 0.1% prevalence) using the IC transmission model. To obtain this ratio, the relationship of IC model-predicted SMA incidence with prevalence was smoothed using LOESS methods.

We calibrated the model for each geographic area so that the total hospitalised SMA incidence was equal to the Paton *et al* estimate. We did this by varying the incidence of SMA in the low-risk children in the *G* state using the *optim* function in R.[16] Given uncertainties about the probability of accessing hospital care, we varied the assumed percentage of  cases eligible for hospitalisation who would have accessed hospital care in the Paton *et al*. settings between 30, 50 and 70%. Likewise we then varied the proportion of cases accessing hospital care in all countries from 30-70%, so that the incidence of hospitalised SMA could be lower, the same, or higher than the Paton *et al* estimates for a given parasite prevalence (from 0.4-fold lower to 2.3-fold higher). We assumed that 50% of uncomplicated malaria episodes would have been treated in the settings studied in Paton et al, based on household survey data.[17] We ran the model in the presence and absence of PDMC for each subnational area and estimated PDMC impact by comparing the two outputs.

### Supplementary Figures

Figure S1. Drug protection against reinfection over time in placebo and PDMC trial arms in fully drug-adherent patients, using drug protection profiles estimated previously.[1,5] Left: placebo arm individuals receive one course of AL at discharge i.e. day 0. Right: PDMC arm individuals additionally receive three courses of DP beginning at days 14, 42 and 70 post-discharge. Neither arm receives any further chemoprevention for the last months of follow up.

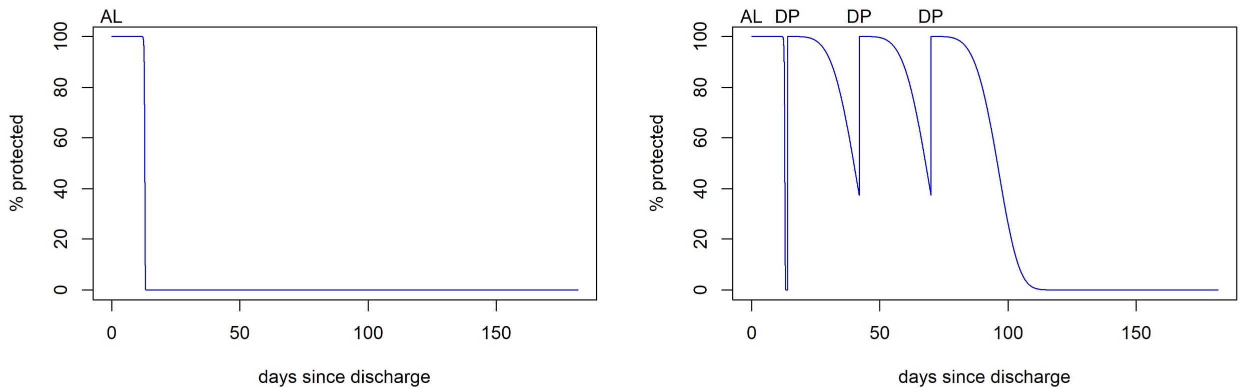

Figure S2. Incidence of hospitalised SMA per 1000 person-years. Incidence in 3 month-9 year olds is plotted as in an original publication by Paton *et al.* Incidence in 0-5 year olds was estimated using the Paton *et al.* result combined with the relative incidence of SMA by age in the IC model (see supplementary text for details).

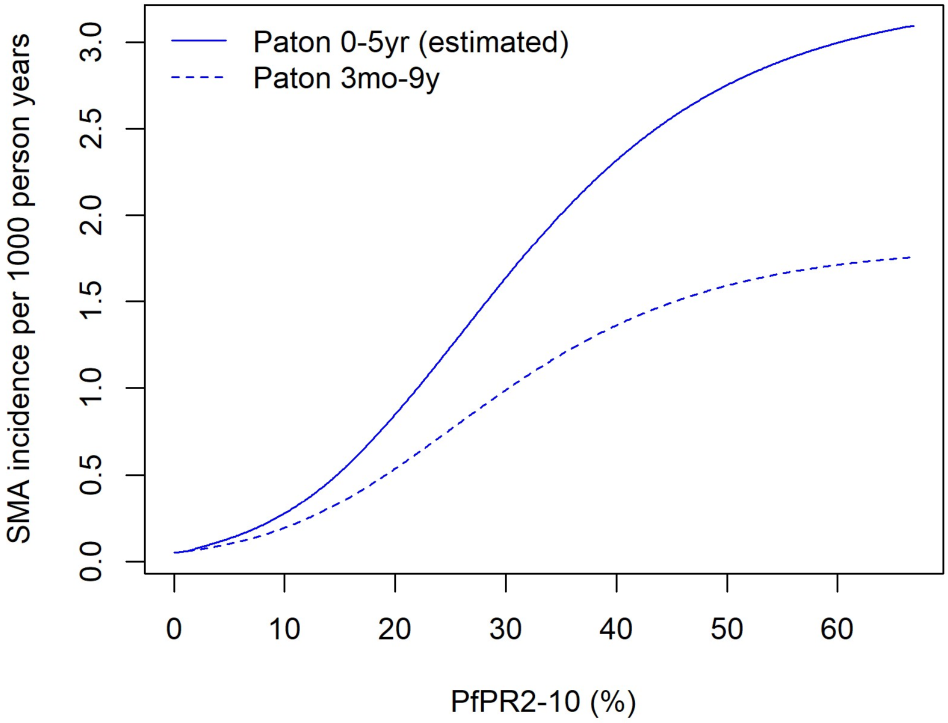

Figure S3. Population model of SMA in all under five year olds.

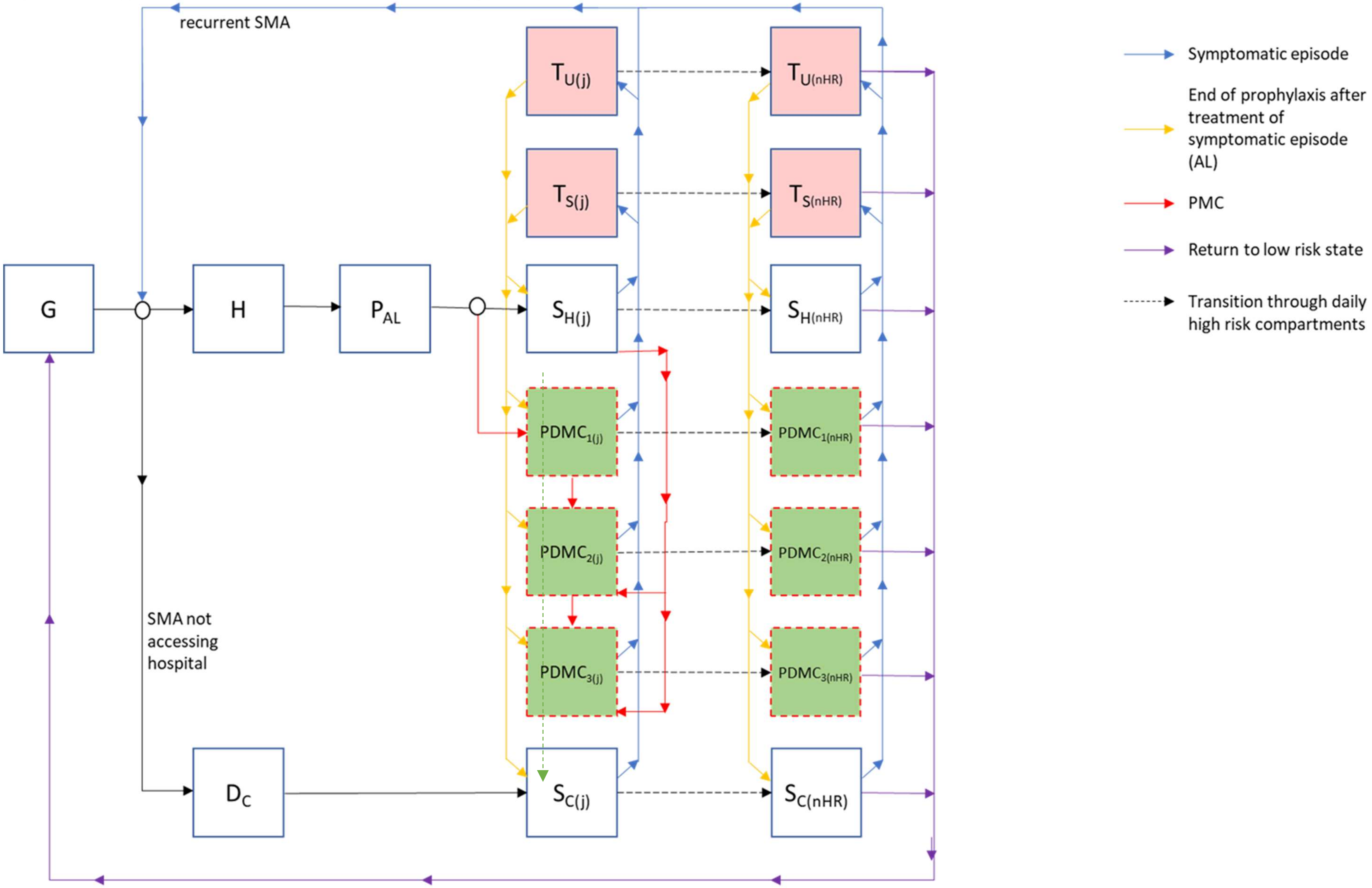

Figure S4. The incidence of symptomatic malaria (uncomplicated & severe) declines over time since discharge. We plot the incidence of symptomatic malaria in the post-discharge cohort relative to EIR in adults (the term $b\phi\xi$ in Table S1, i.e. the probability that an infectious bite leads to infection $b$ , the probability of developing symptoms of clinical disease (total uncomplicated and severe) $\phi$, and the relative exposure to mosquito bites among the post-discharge group of children $\xi$ ).

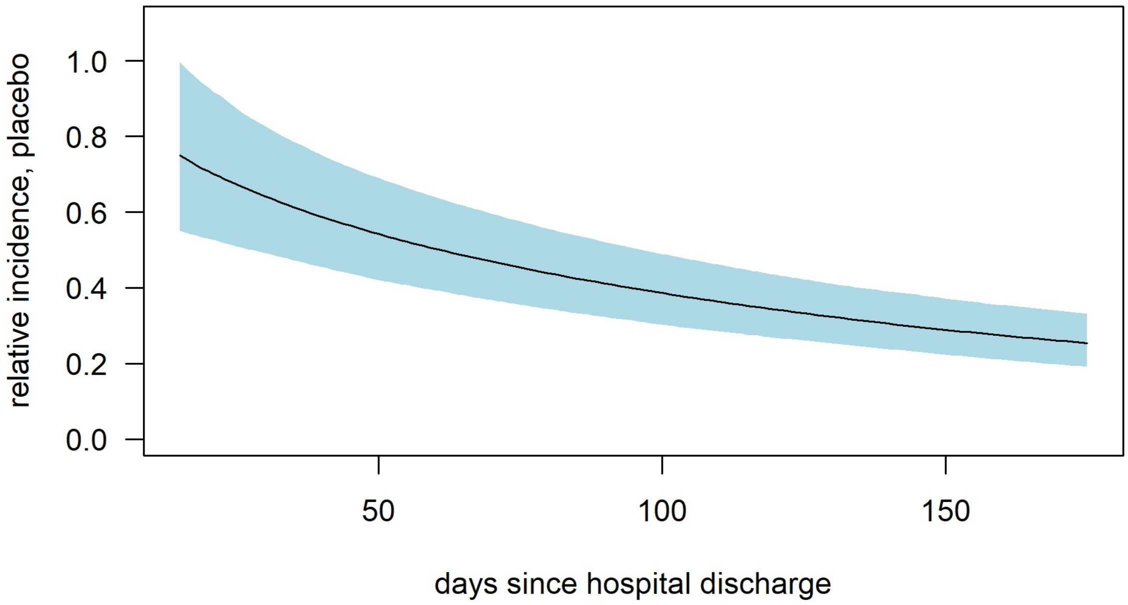

Figure S5. The incidence of symptomatic malaria (uncomplicated & severe) increases with EIR but the incidence per infectious bite is estimated to decline as EIR increases. We plot the relative incidence of symptomatic malaria per bite in the post-discharge cohort relative to EIR in adults (the term $b\phi\xi$ in Table S1, i.e. the probability that an infectious bite leads to infection $b$ , the probability of developing symptoms of clinical disease (total uncomplicated and severe) $\phi$ and the relative exposure to mosquito bites among the post-discharge group of children $\xi$ ).

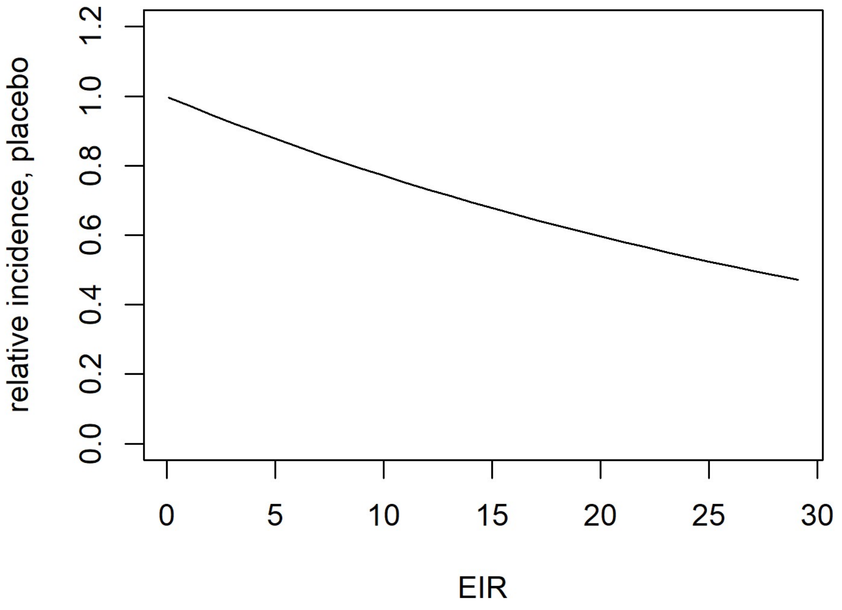

Figure S6: EIR in 0-5-year olds versus observed total incidence of symptomatic malaria in 0-5 year old children per person per year in post-discharge studies, without PDMC. EIR in 0-5-year olds is estimated to be 36% of that experienced by adults, as established in previous studies.[3,18] Symptomatic malaria includes both uncomplicated (UM) and severe malaria (SM). Data are from the 9 trial hospitals in Kwambai *et al*.[4] (K) and validation studies by Opoka *et al*.(O) [19,20] described in the main text. The dashed line indicates the expected relationship if each bite led to a symptomatic malaria episode, and children who have recently had SMA have the same exposure to infectious bites as the average child in the 0-5 year age group. Usually only a fraction of infectious bites results in an infection in the body, and only a fraction of these lead to symptoms.[3,21] The observed relationship suggests children experiencing SMA are more highly exposed to bites than the average child, and/or are more likely to become infected when bitten and experience symptoms.

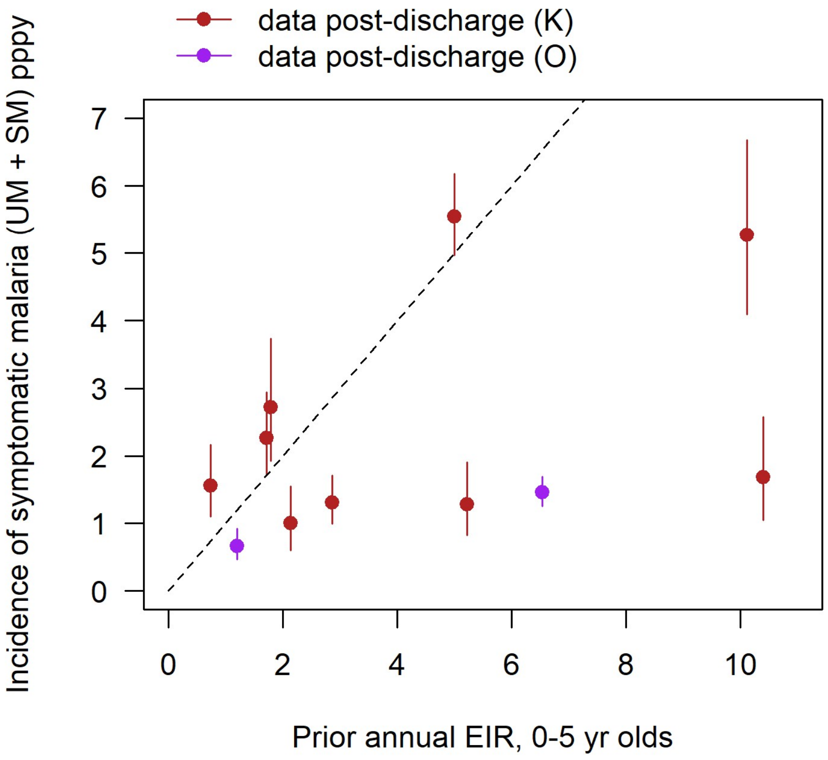

Figure S7. Cumulative number of uncomplicated and hospitalised malaria cases over time since initial hospital discharge in the PDMC trial. The upper lines in each plot show cases in the placebo group, while the lower lines are cases in the PDMC intervention group. The final week highlighted in orange is an outlier with an unexpected number of uncomplicated cases in both trial and placebo arms. These higher numbers occurred just in one trial site in Jinja. We fit the model to incidence in weeks 1-25, excluding the final week 26.

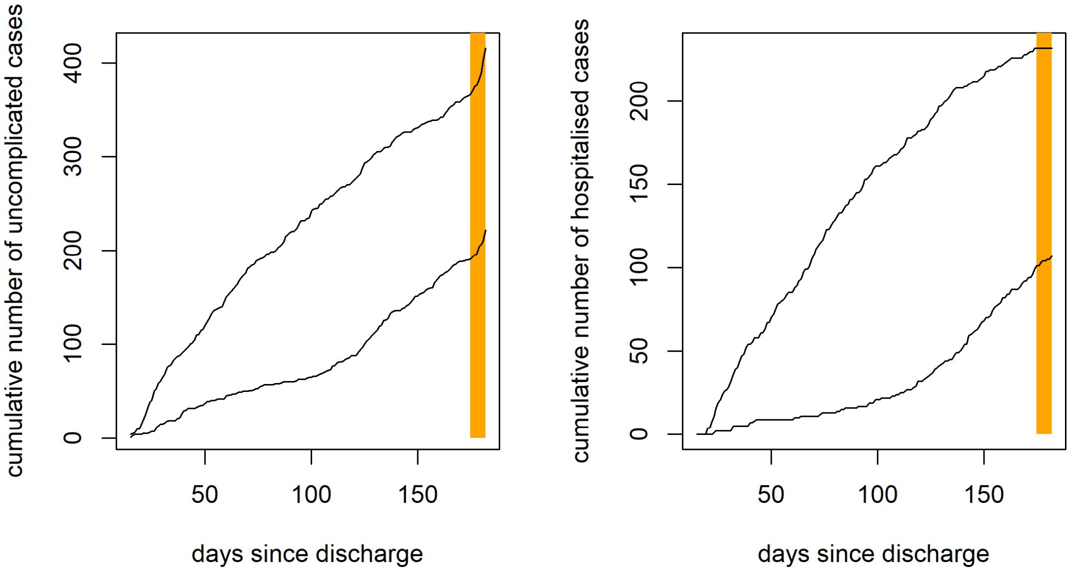

Figure S8. Sensitivity analysis of main text Figure 3C: varying the proportion of cases hospitalised. Assumptions are: (A) 30% of SMA cases were hospitalised in the Paton *et al.* study, to which the baseline incidence of SMA in this model was calibrated, and 70% of cases are hospitalised in the currently modelled setting; (B) 50% of cases hospitalised in Paton *et al.* and 50% in current model, as in main text Figure 4C; (C) 70% of cases hospitalised in Paton *et al.* and 30% in current model. The total number of cases changes but the relative contribution of post-discharge cases to total disease burden in the absence of PDMC remains similar. PDMC has most impact in the scenario where we assume a low proportion hospitalised in Paton *et al.* and a higher proportion in the current model.

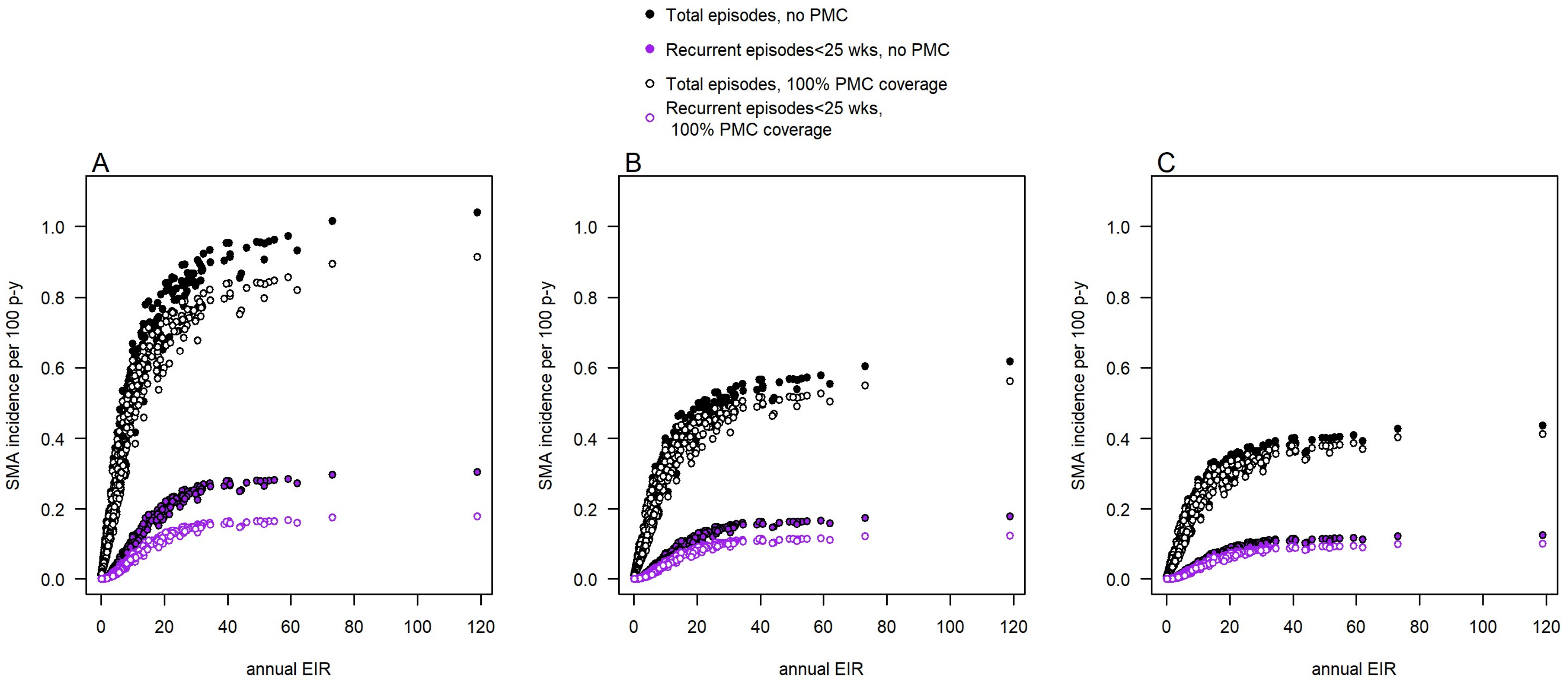

Figure S9. Forecast PDMC demand: the estimated number of treatments required per year per 100 children aged 0-5 years in subnational (admin-1) regions of malaria-endemic countries. Calculations based on incidence of hospitalised SMA in 0–5-year-old children, assuming a 7.4% in-hospital case fatality rate. Panels A, B and C show the lowest, base scenario and highest forecasts of demand, respectively, based on varying assumptions about the proportion of patients with SMA that access hospital.

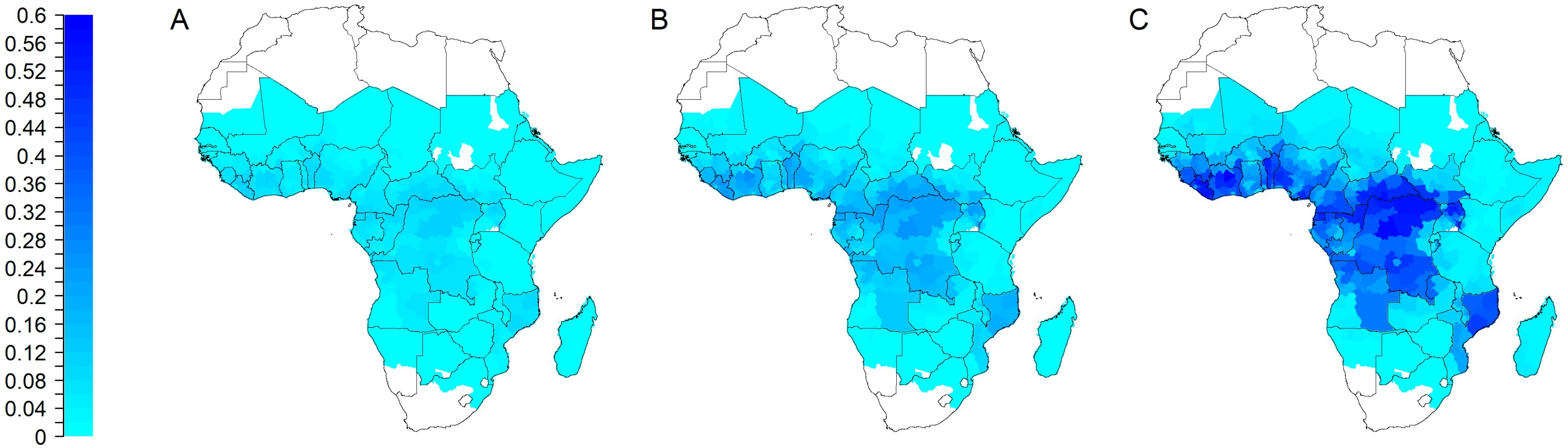

Table S1: Model parameters: post discharge cohort model.

| Parameter | Description | Prior | Posterior value (95% CrI) or fixed value | Source |
|---|---|---|---|---|
| $EIR$ | Annual EIR in adults for each trial site<br><br>Siaya<br>Kisumu<br>Homa Bay<br>Migori<br>Jinja<br>Hoima<br>Masaka<br>Mubende<br>Kamuli | gamma median (95% CI)<br><br>28.0 (22.8-33.8)<br>5.9 (4.8-7.1)<br>14.5 (11.8-17.5)<br>2.0 (1.7-2.4)<br>13.8 (11.3-16.7)<br>7.9 (6.5-9.6)<br>4.7 (3.9-5.7)<br>4.9 (4.0-6.0)<br>28.8 (23.5-34.8) | <br><br><br>28.4 (23.7, 34.2)<br>5.4 (4.5, 6.4)<br>10.9 (8.7, 13.3)<br>2.5 (2.1, 2.9)<br>18.5 (16.0, 21.2)<br>5.5 (4.5, 6.6)<br>5.2 (4.3, 6.1)<br>5.3 (4.5, 6.4)<br>24.5 (18.1, 34.5) | Source of prior means [10,13] |
| $b$ | Probability of successful mosquito-to-human transmission per infectious bite in post-discharge period (episodes per infectious bite) | - | Estimated as part of the product $b\phi\xi$ | - |
| $\phi$ | Proportion of new infections that are symptomatic | - | Estimated as part of the product $b\phi\xi$ | - |
| $\xi$ | Exposure to infectious bites in post-discharge cohort relative to EIR in adults | - | Estimated as part of the product $b\phi\xi$ | - |
| $b\phi\xi$ | Maximum relative incidence of symptomatic malaria in post discharge cohort relative to EIR in adults | Uniform (0,2) | 1.00 (0.61, 1.84) | This analysis |
| $w$ | Rate parameter for the relationship of EIR with probability of symptomatic malaria post-discharge | Half normal (0.0, 1.0) | 0.026 (0.011, 0.040) | This analysis |
| $\theta_1$ | Probability of requiring hospitalisation post-discharge among symptomatic malaria cases, not under active PDMC protection* | Uniform (0,1) | 0.39 (0.36, 0.42) | This analysis |

| Parameter | Description | Prior | Posterior value (95% CrI) or fixed value | Source |
|---|---|---|---|---|
| $\theta_2$ | Probability of requiring hospitalisation post-discharge among symptomatic malaria cases, under active PDMC protection* | Uniform (0,1) | 0.25 (0.17, 0.34) | This analysis |
| $\lambda_{risk}$ | Scale parameter describing the decline in incidence over time post discharge | Half normal (0, 500) | 108.4 (27.3, 186.1) | This analysis |
| $\eta_{risk}$ | Shape parameter describing the decline in incidence over time post discharge | Half normal (0, 500) | 0.64 (0.34, 1.29) | This analysis |
| $f_T$ | Proportion of symptomatic children who access antimalarial treatment | fixed | 1 (in post discharge trial) | Assumption |
| $1/r_{UM}$ | Days until treatment sought after developing first symptoms of malaria | Fixed | 2 | |
| $e_{AL}$ | Efficacy of AL at clearing parasites | fixed | 0.98 | [22] |
| $e_{DP}$ | Efficacy of DP at clearing parasites | fixed | 0.99 | [22] |
| $dur_{TU}$ | Duration in state $T_U$, days | fixed | 15 | [1,23] |
| $dur_{TS}$ | Duration in state $T_S$, days | fixed | 18 | [1,4,23] |
| $dur_{PAL}$ | Duration of post-treatment prophylaxis provided by AL | fixed | 13 | [1] |
| $dur_H$ | Duration of time in hospital for severe malaria, days | fixed | 3 | [4] |
| | Adherence: proportion taking each course of PDMC drugs in the trial setting | | | [4] |
| $p_{ad1}$ | 1st course | fixed | 1 | |
| $p_{ad2}$ | 2nd course | fixed | 0.985 | |
| $p_{ad3}$ | 3rd course | fixed | 0.973 | |

| Parameter | Description | Prior | Posterior value (95% CrI) or fixed value | Source |
|---|---|---|---|---|
| | Duration of protection after AL treatment, gamma-distributed | | | [1] |
| $k_{AL}$ | Shape parameter | fixed | 93.5 | |
| $v_{AL}$ | Scale parameter | fixed | 0.139 | |
| | Duration of protection after DP treatment, Weibull-distributed | | | [5] |
| $\lambda_{DP}$ | Scale parameter | fixed | 28.1 | |
| $\eta_{DP}$ | Shape parameter | fixed | 4.4 | |
| $inc_d$ | Incidence of symptomatic episodes of malaria per person per day (uncomplicated + more severe) post-discharge | variable | | - |
| $y_d(t_d)$ | Probability of a symptomatic malaria episode per person per day (uncomplicated + more severe) at time $t_d$ post-discharge | variable | | - |
| $p_{protectDP}$ | Probability of protection against re-infection by DP by day since treatment | fixed, dependent on time since treatment | | - |

Table S2: Additional model variables and parameters used in the full population model of SMA that are not already covered in Table S1.

| Variable or parameter | Description | Value | Source |
|---|---|---|---|
| $C_{SMA}(t)$ | Number of new SMA cases on day *t* | Variable | current analysis |
| $C_S(t)$ | Number of new, severe non-SMA cases on day *t* (cases eligible for hospitalisation) in the high risk post-SMA states | Variable | current analysis |
| $y_{gSMA}$ | Probability of a symptomatic malaria episode per person per day (uncomplicated + severe) in the *G* state | Variable | current analysis |
| $n_{HR}$ | Number of days in the high risk state from weeks 3-25 after discharge | 161 | current analysis |
| $p_{SMA}$ | Probability of having SMA among severe episodes post-discharge | 0.44 | [4] |
| $p_H$ | Probability of severe cases accessing hospital care | Varied: 0.3, 0.5, 0.7 | [14] |
| $z_{PMC}$ | PDMC coverage: proportion of children given PDMC at hospital discharge | 1 | Assumed |
| $f_T$ | Proportion of symptomatic children who access antimalarial treatment | 0.5<br>Outside trial settings | [17] |
| $q_{SMAH}$ | Case fatality rate: hospitalised SMA | 7.4% | [24] |
| $q_{SMAC}$ | Case fatality rate: non-hospitalised SMA | 14.8% | derived from [24,25] |
| $q_H$ | Case fatality rate: hospitalised non SMA severe malaria | 1.0% | [25] |
| $q_C$ | Case fatality rate: severe malaria non-hospitalised | 2.0% | derived from [25] |
| $p_{ad}$ | Adherence: proportion taking all 3 doses of each course of PDMC drugs during routine implementation<br>1st course (day 14)<br>2nd course (day 42)<br>3rd course (day 70) | <br><br><br><br>0.765<br>0.879<br>0.894 | [15] |
